# Mindfulness-Based Stress Reduction for Breast Cancer Survivors (MBSR (BC)): Evaluating Mediators of Psychological and Physical Outcomes in a Large Randomized Controlled Trial

**DOI:** 10.1101/2020.05.05.20092171

**Authors:** Cecile A. Lengacher, L. Forest Gruss, Kevin E. Kip, Richard R. Reich, Manolete S. Moscoso, Katterine G. Chauca, Anisha Joshi, Pinky Budhrani Shani, Lakeshia Cousin, Carly Paterson Khan, Matthew Goodman, Jong Y. Park

**Author notes:** **Address for Correspondence:** Cecile A. Lengacher, University of South Florida, College of Nursing, MDC 22, 12901 Bruce B. Downs Blvd., Tampa, FL 33612-4476., (email). **Data Repository:** Data will be archived in accordance with the NIH Data Sharing Plan, a national data repository.

## Abstract

MBSR(BC) is known to have a positive impact on psychological and physical symptoms among breast cancer survivors (BCS). However, the cognitive mechanisms of “how” MBSR(BC) works are unknown. The purpose of this study, as part of a larger R01 trial, was to test whether positive effects achieved from the MBSR(BC) program were mediated through changes in increased mindfulness, decreased fear of breast cancer recurrence, and perceived stress. Female BCS >21 years diagnosed with Stage 0-III breast cancer were randomly assigned to a 6-week MBSR(BC) or a Usual Care(UC) regimen. Potential outcome mediators were identified by use of an analysis of covariance (ANCOVA), comparing mean values of outcome variables and potential mediating variables followed by mediational and bootstrap analyses. Among 322 BCS (167 MBSR(BC) and 155 UC), fear of recurrence and perceived stress, but not mindfulness, mediated reductions in anxiety and fatigue at weeks 6 and 12, partially supporting our hypothesis of cognitive mechanisms of MBSR(BC).

**Support:** This study was supported by the National Cancer Institute (Award Number 1R01 CA131080-01A2). This work also has been supported in part by the Biostatistics and Bioinformatics Shared Resource at the H. Lee Moffitt Cancer Center & Research Institute, an NCI designated Comprehensive Cancer Center (P30-CA076292). The content is solely the responsibility of the authors and does not necessarily represent the official views of the National Cancer Institute or the National Institutes of Health. This study protocol was approved by the Institutional Review Board at the University of South Florida to ensure the ethical treatment of participants.

**Conflict of Interest:** The authors have no conflicts to report.

**Trial Registration:** www.ClinicalTrials.gov Registration Number: NCT01177124

## INTRODUCTION

Breast cancer is the most prevalent cancer among women, with 3.8 million cases reported in the United States for 2019 (DeSantis et al., 2019). Although 90% of breast cancer survivors (BCS) are surviving at least five years (Society A.C., 2020) after diagnosis due to new advanced treatments, they are often unprepared for the associated emotional trauma and multiple psychological and physical symptoms impacting their quality of life (QOL)(DeSantis et al., 2019; C. A. Lengacher et al., 2014; Society, 2019). Evidence shows Mindfulness-based stress reduction (MBSR) programs offer a safe non-pharmacological approach for BCS providing multiple health benefits to reduce cancer symptom burden while also increasing their QOL (C. A. Lengacher, et al., 2016). Specific evidence shows mindfulness-based programs are associated with improvements in psychological health, (stress, anxiety and depression) (C. A. Lengacher et al., 2012), and physical symptoms, (pain, fatigue and sleep) (Carlson et al., 2013; Garland et al., 2014; Johns et al., 2015; C. A. Lengacher et al., 2015; C. A. Lengacher, Reich, Paterson, Ramesar, Park, Alinat, Johnson-Mallard, Moscoso, Budhrani-Shani, et al., 2016), yet there is limited research examining the “mechanism of action” of mindfulness training to provide insight into the process by which this training impacts patient outcomes. Research utilizing mediation analyses may advance the scientific basis of how behavioral interventions work (Alan E Kazdin, 2006; Moyer et al., 2012), by also validating effective components on individualized patient benefits (Laurenceau, Hayes, & Feldman, 2007).

The “healing power” of mindfulness is postulated to occur through two cognitive processes: awareness and attention (Brown & Cordon, 2009). Training in meditation is proposed to increase self-regulation of emotions and thoughts (Bishop, 2002), through use of moment-to-moment, non-judgmental/non-reactive awareness to internal and external experiences, resulting in reduced rumination and elaboration over past or future distressing experiences (Baer, 2003; Bishop et al., 2004; K. W. Brown, R. A. Ryan, & J. D. Creswell, 2007; Kabat-Zinn, 1990, 2003). The mechanisms by which MBSR(BCS) improves health outcomes/well-being remain largely unknown and mechanisms have not been validated (Moyer et al., 2012). Primary targets of mechanisms have been mindfulness, in addition to emotion regulation and cognitive control in the form of altering one’s perspective as a function of mindfulness practice.

For BCS, evidence shows MBSR significantly reduces perceived stress, anxiety and fatigue (C. A. Lengacher, Reich, Paterson, Ramesar, Park, Alinat, Johnson-Mallard, Moscoso, Budrhani Shani, et al., 2016; Reich et al., 2017; Zainal, Booth, & Huppert, 2013). Mindfulness has been examined not just as an outcome measure, but as a mediator, and therefore a potential mechanism of MBSR among cancer survivors. When examining mindfulness as a mediator of stress, improvements in perceived stress were found among survivors with a past diagnosis of cancer (Branstrom, Kvillemo, Brandberg, & Moskowitz, 2010) and specifically among BCS (Boyle, Stanton, Ganz, Crespi, & Bower, 2017). While previous research investigated perceived stress as an outcome, there continues to be limited evidence investigating “stress” as a mechanism that modifies health outcomes. In a recent study among gastrointestinal cancer patients, perceived stress was found to mediate the relationship between dispositional mindfulness and psychological symptoms (anxiety, depression, social dysfunction and loss of confidence) (Xu, Zhou, Fu, & Rodriguez, 2017).

In addition to mindfulness and perceived stress, fear of recurrence has been debated as a mechanism by which the MBSR intervention exerts positive outcomes on psychological and physical health among cancer survivors. Often for breast cancer survivors, worry, rumination and uncertainty underlie the *symptom expression* of fear of cancer recurrence (Lee-Jones, Humphris, Dixon, & Hatcher, 1997). In addition, “intolerance of uncertainty” has been identified to be positively related to cancer-related distress in prostate cancer patients (Eisenberg et al., 2015). Cancer survivors who expressed a clinical fear of cancer recurrence reported a high degree of “intolerance of uncertainty” related to cancer recurrence (Mutsaers et al., 2016). Women with breast and ovarian cancer showed reduced fear of recurrence, along with reduced cancer-specific uncertainty after psychotherapeutic intervention (Lebel et al., 2014). MBSR reduced cancer-related anxiety and intolerance of uncertainty, and increased mindfulness, among prostate cancer patients up to 12-months post diagnosis (Victorson et al., 2012). MBSR(BC) and other mindfulness interventions have also reduced fear of cancer recurrence while concurrently enhancing mindfulness and self-regulation (Butow et al., 2017; Cheli et al., 2019; Compen et al., 2018; Crane-Okada et al., 2012; C.A. Lengacher et al., 2011; C. A. Lengacher et al., 2009; C. A. Lengacher, Reich, Paterson, Ramesar, Park, Alinat, Johnson-Mallard, Moscoso, Budrhani Shani, et al., 2016). Evidence is lacking in that fear of recurrence may also function as a mediator of reduced physical symptoms, such as fatigue, a common symptom reported by BCS transitioning off treatment.

Our first mediation study, among 82 breast cancer survivors who received MBSR(BC) versus UC (C. A. Lengacher et al., 2014), showed that a reduction in fear of recurrence and improvement in physical functioning mediated improvements in perceived stress and anxiety. These findings suggest that MBSR(BC) may impart “self-regulated” changes in cognitive and affective appraisals of one’s experiences and perspectives. Improved mindfulness and enhanced emotion regulation through MBSR have resulted in reduced mood disturbances (Laura E Labelle, Lawlor-Savage, Campbell, Faris, & Carlson, 2015). In this study, we hypothesize cognitive mechanisms are mediators of the MBSR(BC) intervention for improving psychological health, through the processes of self-regulation of emotions to internal and external experiences. Our first mediation study (C. A. Lengacher et al., 2014), was limited to investigating fear of recurrence as a potential cognitive mechanism through which symptom improvements may have occurred. This current R01 investigated “mindfulness” and “perceived stress” as additional mediators of the MBSR(BC) intervention among BCS.

The aim of this study, as part of a larger R01 trial, was to test for potential mediators of MBSR(BC) on symptom improvement to provide insight into the bio-behavioral “mechanisms of action,” or “how” the MBSR(BC) program works to yield its salutatory effects.

## METHODS

### Sample and Setting

Female breast cancer survivors (BCS; n=322) aged 21 and older were recruited from Moffitt Cancer Center, and the Carol and Frank Morsani Center for Advanced Healthcare between April 2009 through March 2014. BCS were included with a diagnosis of Stage 0-III breast cancer, lumpectomy and/or mastectomy and/or adjuvant radiation and/or chemotherapy. BCS with a diagnosis of Stage IV cancer, a severe mental disorder, and/or breast cancer recurrence were excluded.

### Study Design and Randomization

BCS were randomly assigned in a 1:1 ratio to either: (i) the formal (in-class) 6-week MBSR(BC) program tailored to BCS; or the (ii) usual care (UC) waitlisted regimen in which MBSR(BC) was offered within 6 months of enrollment. Participants were stratified by type of surgery (lumpectomy versus mastectomy), breast cancer treatment (chemotherapy with or without radiation versus radiation alone), and stage of breast cancer (Stage 0/I versus II/III).

### Procedures

The Institutional Review Board at the University of South Florida approved all procedures. BCS who met study inclusion criteria and expressed interest in the study were invited to an orientation session. If they agreed to participate, informed consent was obtained, and baseline data were collected on measures of psychological and physical symptom status, QOL, demographic characteristics, and clinical history followed by randomization to either the MBSR(BC) program or the UC waitlisted group. BCS completed assessments at 6 weeks upon program completion and at 12 weeks, six weeks after program completion. The UC group attended standard post-treatment clinic visits and were wait-listed to receive the MBSR(BC) program after the 12-week study period.

### MBSR(BC) intervention procedures

BCS randomized to the MBSR(BC) intervention attended 6 weekly (2-hour) sessions conducted by a trained psychologist in MBSR(BC). BCS received a training manual and 5 audio CD’s to guide their home practice of sitting meditation, body scan, gentle yoga, and walking meditation and recorded practice time in a diary during the 12 weeks of the study. The MBSR(BC) program is a 6-week program adapted by Dr. Lengacher for BCS from Jon Kabat-Zinn’s 8-week program (Kabat-Zinn, Lipworth, & Burney, 1985; Kabat-Zinn et al., 1992). The intervention consists of: 1) educational materials; 2) practice sessions of 4 formal and informal meditative techniques; and 3) group processes related to barriers to the practice, application in daily situations, and supportive group interaction. Participants receive meditative training (Kabat-Zinn et al., 1992) in 4 formal meditation techniques: 1) sitting meditation; 2) body scan meditation; 3) Gentle Hatha Yoga; and 4) walking meditation. BCS are taught to bring awareness to thoughts and emotions associated with symptoms such as pain, anxiety, sleep disturbances, and fear of recurrence and to then separate the emotional experience from the sensory experience. Through MBSR(BC), BCS learn to mitigate fears attached to physical and emotional distress by promoting self-regulation of attention through meditative practices. Informal techniques of mindfulness are learned by integrating attention and awareness into their daily activities (Hamilton, Kitzman, & Guyotte, 2006). An example of self-regulation is the process of not getting caught up, or ruminating over something unpleasant, such as pain or anxiety, but focusing attention on the breath and the current activity. As one is immersed in the task, calmness occurs; this is called “present moment awareness,” a central concept of MBSR (Hamilton et al., 2006).

## Measures

### Physical Symptoms

Physical symptoms included the following measurements. ***Pain***. The Brief Pain Inventory (BPI) measures pain, including intensity and interference, and has reliability coefficients ranging from 0.82 to 0.95 (Keller et al., 2004). ***Fatigue***. The Fatigue Symptom Inventory (FSI) is a 14-item self-report questionnaire that measures fatigue severity, frequency, daily pattern, and perceived interference with QOL (Hann, Jacobsen, Martin, Azzarello, & Greenberg, 1998). The FSI has a reliability alpha coefficient of 0.90, and a test-retest reliability ranging from r=.35-.75 (Keller et al., 2004). ***Sleep***. The Pittsburgh Sleep Quality Index (PSQI) is a 19 self-report sleep questionnaire with 5 questions rated by the bed partner to rate sleep quality of patients. Reliability is 0.80 for the global PSQI and ranges from 0.70 to 0.78 for sleep disturbance (Carpenter & Andrykowski, 1998).

### Psychological symptoms

Psychological symptoms included the following measurements. ***Depression***. The Center for Epidemiological Studies Depression Scale (CES-D), a four-point scale evaluated depression and has a reliability coefficient of 0.92 for breast cancer subjects (Radloff, 1977). ***State anxiety***. The State Trait Anxiety Inventory measured anxiety; internal consistency reliability is reported as 0.95 (Spielberger, Gorsuch, & Luschene, 1983). ***Perceived stress***. The Perceived Stress Scale (PSS) measured participant stress; it is a 14-item Likert-type scale with an internal consistency reliability ranging from 0.84 to 0.86 (Cohen, Kamarck, & Mermelstein, 1983). ***Fear of cancer recurrence***. The Concerns about Recurrence Scale measured fear of recurrence (cancer); it is a 30-item scale including four items assessing ‘‘worry’’ related to recurrence, scored as (1) ‘‘I don’t think about it at all’’ to (6) ‘‘I think about it all the time” and 26 items assessing the nature of the fear of recurrence and extent to which they worry about each item, scored on a five-point Likert scale ranging from (0) not at all, to (4) extremely. Overall, internal consistency reliability is 0.87 for BCS (Vickberg, 2003). ***Mindfulness*** was measured by 2 instruments. The Five Facet Mindfulness Questionnaire (FFMQ) (Baer, Smith, Hopkins, Krietemeyer, & Toney, 2006), a 39-item instrument on a 1-5 Likert Scale evaluated five factors of mindfulness, observing, describing, acting with awareness, non-judging of inner experience, and non-reactivity to inner experience. Internal consistency ranges from 0.72 to 0.92 (Baer et al., 2008). The Cognitive and Affective Mindfulness Scale-Revised (CAMS-R) (Feldman, 2007), is a 12-item 1-4 Likert-scale measuring 4 subscales of mindfulness (attention, present-focus, awareness, acceptance/non-judgment); internal consistency ranges from 0.74 to 0.77. **Quality of Life (QOL)**. The Medical Outcomes Study Short Form was used to assess mental and physical health as related to QOL; higher scores are demonstrative of better mental and physical health. Health-related quality of life was measured by the Medical Outcomes Studies Short-form (MOS SF-36), a 36-item questionnaire with Likert-type responses. Internal consistency reliability ranged from 0.62 to 0.94 (Ware, Kosinski, & Keller, 1994). Test-retest reliability ranged from 0.43 to 0.90 (Ware et al., 1994). **Demographic data/clinical history**. Standard socio-economic demographic data were collected on age, gender, ethnicity, education completed, marital status, income status, and employment status at baseline and updated at 6 and 12 weeks. Standard clinical history data were collected at baseline, 6 and 12 weeks and included type of treatment, cancer diagnosis, and dates on treatment. As part of the clinical history form, data included a social history, lifestyle health behaviors, and medication use.

### Statistical methods

Baseline demographic/clinical characteristics of outcomes and potential mediators were summarized as means and standard deviations for continuous variables and percentages for categorical variables. Patient characteristics were compared by random assignment by use of student *t* tests for continuous variables and chi-square tests for categorical variables. To evaluate whether positive effects achieved from the MBSR(BC) program were mediated through changes in mindfulness, perceived stress, and fear of recurrence of breast cancer: Analysis of covariance (ANCOVA) compared mean values of primary outcome variables and potential mediating variables at 6 and 12-weeks by random assignment and were adjusted for baseline values. On the basis of this analysis, outcome variables and potential mediating variables showing the strongest main effects associated with MBSR(BC) were selected for the mediational analyses. The meditational analyses were conducted using the methods described by Preacher and Hayes (2004) and Imai et al. (2010) with *p*-values determined by use of bias corrected bootstrap confidence intervals. The percent of total effects explained by the mediator, were calculated as described by Hicks and Tingley (2011). With this method, estimates of total effects of MBSR(BC) on the primary outcomes of interest, and indirect effects attributed to a potential mediator, were estimated and plotted. These analyses included baseline to 6-week measurements and baseline to 12-week measurements. The sequential assumption common in mediation analysis (Imai et al., 2010) was not tested because treatment was randomized. The SAS System, version 9.4 (Cary, NC) was used for analyses, with a 2-sided p-value of 0.05 used to define statistical significance.

## RESULTS

### Participant characteristics

Among the 322 BCS enrolled, 299 completed the study 167 MBSR(BC) and 155 UC participants (C. A. Lengacher, et al., 2016) for the CONSORT chart reporting number of participants screened, randomized, and retained, as well as demographic and clinical characteristics by random assignment.

Results of the calculated baseline values of the primary outcome and potential mediators of interest by random assignment are presented in Table 1. In contrast to baseline demographic and clinical characteristics, several of the *primary outcome* and potential *mediating variables* were not fully balanced by random assignment. This included the MBSR(BC) intervention group compared to the UC group as having significantly higher state anxiety, fatigue, fear of recurrence overall, and fear of recurrence problems, and perceived stress at baseline, and lower QOL (emotional well-being). Mean scores for the two measures of mindfulness were similar by random assignment.

**Table 1.** Primary Outcome and Potential Mediating Variables at Baseline by Random Assignment*.

| **Outcome Variable** | **All (n=320)** | **UC (n=155)** | **MBSR (n=165)\*\*** | **p-value** |
| --- | --- | --- | --- | --- |
| Depression (CES-D) | 10.5 (6.7) | 10.0 (6.5) | 10.9 (6.9) | 0.27 |
| State Anxiety (STAI) | 37.3 (11.9) | 35.9 (11.3) | 38.6 (12.3) | 0.04 |
| Fatigue (FSI) | 15.4 (8.6) | 14.5 (8.4) | 16.4 (8.8) | 0.05 |
| Sleep Disturbance (PSQI) | 1.8 (0.7) | 1.8 (0.6) | 1.8 (0.7) | 0.86 |
| Pain severity (BPI) | 10.6 (9.4) | 9.7 (8.6) | 11.4 (10.1) | 0.10 |
| Physical functioning (SF-36) | 64.7 (28.1) | 64.7 (28.0) | 64.7 (28.3) | 0.99 |
| Emotional well-being (SF-36) | 66.1 (18.0) | 68.4 (18.3) | 64.0 (17.4) | 0.03 |
| General health (SF-36) | 64.4 (21.1) | 66.3 (20.8) | 62.6 (21.3) | 0.12 |
| **Potential Mediator** |  |  |  |  |
| Fear of recurrence (overall) | 11.5 (5.6) | 10.8 (5.6) | 12.2 (5.6) | 0.02 |
| Fear of recurrence (problems) | 35.8 (24.7) | 31.8 (24.5) | 39.4 (24.4) | 0.006 |
| Mindfulness (CAMS-R) | 34.7 (6.9) | 35.4 (7.2) | 34.0 (6.6) | 0.08 |
| Mindfulness (FFMQ)(n=170) | 133.9 (20.6) | 135.5 (20.5) | 132.4 (20.7) | 0.33 |
| Perceived stress (PSS) | 16.5 (7.7) | 15.4 (7.6) | 17.6 (7.7) | 0.01 |

\*Values are presented as mean ± standard deviation. \*\*2 participants did not have baseline values.

### Early Intervention Effects

Mean change scores from baseline to 6-weeks and baseline to 12 weeks by random assignment are presented in Table 2 for a range of outcome variables and potential mediating variables, and adjustment for the baseline value of the outcome of interest. From baseline to 6-weeks, the MBSR(BC) program was associated with greater reductions in state anxiety (p=0.01) and fatigue (*p*=0.006), as well as higher levels of emotional well-being (*p*=0.04). *<u>Early Mediation effects at 6 weeks.</u>* For potential mediating variables, the ANCOVA revealed the MBSR(BC) program was associated with greater reduction in overall fear of recurrence (*p*=0.03), problems stemming from fear of recurrence (*p*=0.0003), and a trend towards lower perceived stress (*p*=0.10) at 6 weeks. Mindfulness was not a significantly affected by MBSR at 6 weeks (*p*=0.49 for CAMS-R scores; *p*=0.27 for FFMQ scores). Of note, all but one baseline to 6-week outcome measure was in the expected direction of improved symptom status with MBSR(BC).

**Table 2.** Main Effects of MBSR on Outcome Variables and Potential Mediating Variables at 6- and 12-Weeks.

| Outcome Variable | Baseline to 6-Weeks |  |  |  |  |  | Baseline to 12 Weeks |  |  |  |  |  |
| --- | --- | --- | --- | --- | --- | --- | --- | --- | --- | --- | --- | --- |
|  | Mean |  | Adjusted |  | ES* | p-value | Mean |  | Adjusted |  | ES* | p-value |
|  | difference |  | Mean** |  |  |  | difference |  | Mean** |  |  |  |
|  | UC<br>(n=155) | MBSR<br>(n=165) | UC<br>(n=155) | MBSR<br>(n=165) |  |  | UC<br>(n=155) | MBSR<br>(n=165) | UC<br>(n=155) | MBSR<br>(n=165) |  |  |
| Depression (CES-D) | -1.3 (4.7) | -2.5 (6.3) | 9.0 (6.0) | 8.0 (5.5) | 0.22 | 0.08 | -1.1 (5.5) | -2.0 (6.1) | 9.2 (6.8) | 8.5 (6.3) | 0.17 | 0.24 |
| State Anxiety (STAI) | -2.6 (8.8) | -6.4 (11.7) | 34.0<br>(11.5) | 31.3<br>(11.2) | 0.36 | 0.01 | -2.7<br>(10.6) | -5.6 (10.6) | 33.9<br>(13.2) | 31.8<br>(11.4) | 0.27 | 0.06 |
| Fatigue (FSI) | -1.3 (6.5) | -3.7 (6.8) | 13.8 (8.5) | 11.9 (7.6) | 0.36 | 0.006 | -1.4 (7.3) | -3.7 (8.5) | 13.6 (8.7) | 11.9 (8.6) | 0.28 | 0.03 |
| Sleep Disturbance (PSQI) | -0.1 (0.6) | -0.2 (0.6) | 1.7 (0.6) | 1.6 (0.7) | 0.13 | 0.21 | -0.2 (0.6) | -0.2 (0.7) | 1.6 (0.7) | 1.6 (0.7) | 0.11 | 0.28 |
| Pain severity (BPI) | -1.4 (6.4) | -1.6 (7.4) | 9.2 (9.4) | 8.8 (8.2) | 0.02 | 0.60 | -1.0 (7.1) | -2.6 (7.8) | 9.1 (8.4) | 8.1 (9.4) | 0.21 | 0.16 |
| Physical functioning (SF-36) | 3.1 (25.3) | 0.0 (19.1) | 67.6<br>(29.5) | 64.6<br>(29.8) | -0.14 | 0.22 | 0.6 (27.5) | 0.3 (23.9) | 65.5<br>(31.7) | 65.3<br>(30.9) | 0.01 | 0.93 |
| Emotional well-being (SF-36) | 1.3 (16.1) | 6.2 (16.0) | 68.2<br>(18.8) | 71.8<br>(18.0) | 0.30 | 0.04 | 4.4 (15.7) | 6.6 (18.9) | 71.6<br>(19.1) | 72.3<br>(19.4) | 0.12 | 0.70 |
| General health (SF-36) | 2.4 (16.0) | 5.1 (12.9) | 67.1<br>(19.9) | 69.0<br>(21.3) | 0.18 | 0.24 | 1.7 (16.8) | 5.5 (15.3) | 66.6<br>(22.1) | 69.6<br>(21.6) | 0.23 | 0.09 |
| **Potential Mediator** |  |  |  |  |  |  |  |  |  |  |  |  |
| Fear of recurrence-overall | -1.0 (3.7) | -2.4 (4.5) | 10.3 (5.4) | 9.3 (5.3) | 0.33 | 0.03 | -1.4 (5.0) | -2.9 (5.2) | 9.8 (5.8) | 8.9 (5.3) | 0.30 | 0.07 |
| Fear of recurrence-problems | -2.1<br>(17.6) | -10.9<br>(17.1) | 32.8<br>(25.7) | 25.7<br>(24.0) | 0.49 | 0.0003 | -3.7<br>(17.2) | -10.7<br>(18.4) | 30.9<br>(26.0) | 25.6<br>(24.5) | 0.39 | 0.008 |
| Mindfulness (CAMS-R) | 1.4 (5.1) | 1.3 (6.1) | 36.1 (7.5) | 35.7 (7.5) | 0.02 | 0.49 | -0.1<br>(10.6) | 1.5 (9.8) | 34.9<br>(11.4) | 36.0<br>(10.5) | 0.15 | 0.32 |
| Mindfulness<br>(FFMQ)(n=170) | 1.9 (18.5) | 6.1 (24.0) | 136 (25) | 139 (30) | 0.20 | 0.27 | 1.9 (18.5) | 2.0 (19.4) | 136 (24) | 143 (36) | 0.30 | 0.08 |
| Perceived stress (PSS) | -2.1 (6.8) | -4.3 (8.0) | 13.9 (7.7) | 12.7 (6.9) | 0.29 | 0.10 | -2.5 (6.9) | -4.3 (6.7) | 13.5 (8.2) | 12.3 (7.7) | 0.26 | 0.11 |

\*Effect size (ES); positive values represent improvement with MBSR. \*\*Adjusted for value at baseline.

### Late Intervention Effects

From baseline to 12-weeks, the MBSR(BC) program was associated with greater reductions in fatigue (*p*=0.03), as well as trending reductions in state anxiety (*p*=0.06) and QOL (*p*=0.09). *<u>Later Mediation Effects</u>* <u>at 12 weeks</u>. For potential mediating variables, the MBSR(BC) program was associated with greater reduction in fear of recurrence problems (*p*=0.008) at 12 weeks and a trend towards lower perceived stress (*p*=0.11). Mindfulness was trending for the FFMQ scores (*p*=0.08) but was not significantly different for the CAMS-R scores (*p*=0.32). On the basis of these results, state anxiety and fatigue were selected as primary outcome variables, and fear of recurrence (problems) and perceived stress were selected as potential mediating variables for mediational analyses. Although perceived stress was only trending for both early and late effects, mediation effects were nevertheless constant throughout the entire study period and perceived stress was therefore included for further mediation analyses.

### Fear of Recurrence as Mediator

As seen in Figure 1 (top half), a significant portion of the relationship between MBSR(BC) and state anxiety at both 6 (*p*=0.03) and 12-weeks (*p*=0.008) was mediated through reductions in fear of cancer recurrence. At both time points, the reduction in fear of cancer recurrence associated with the MBSR(BC) program explained 37% of the total effects of MBSR(BC) on levels of state anxiety. Similarly, reductions in fear of recurrence associated with the MBSR(BC) program mediated reductions in fatigue at 12-weeks (*p*=0.003), but not 6-weeks (*p*=0.17) and (Figure 1, bottom half).

**Figure 1.**
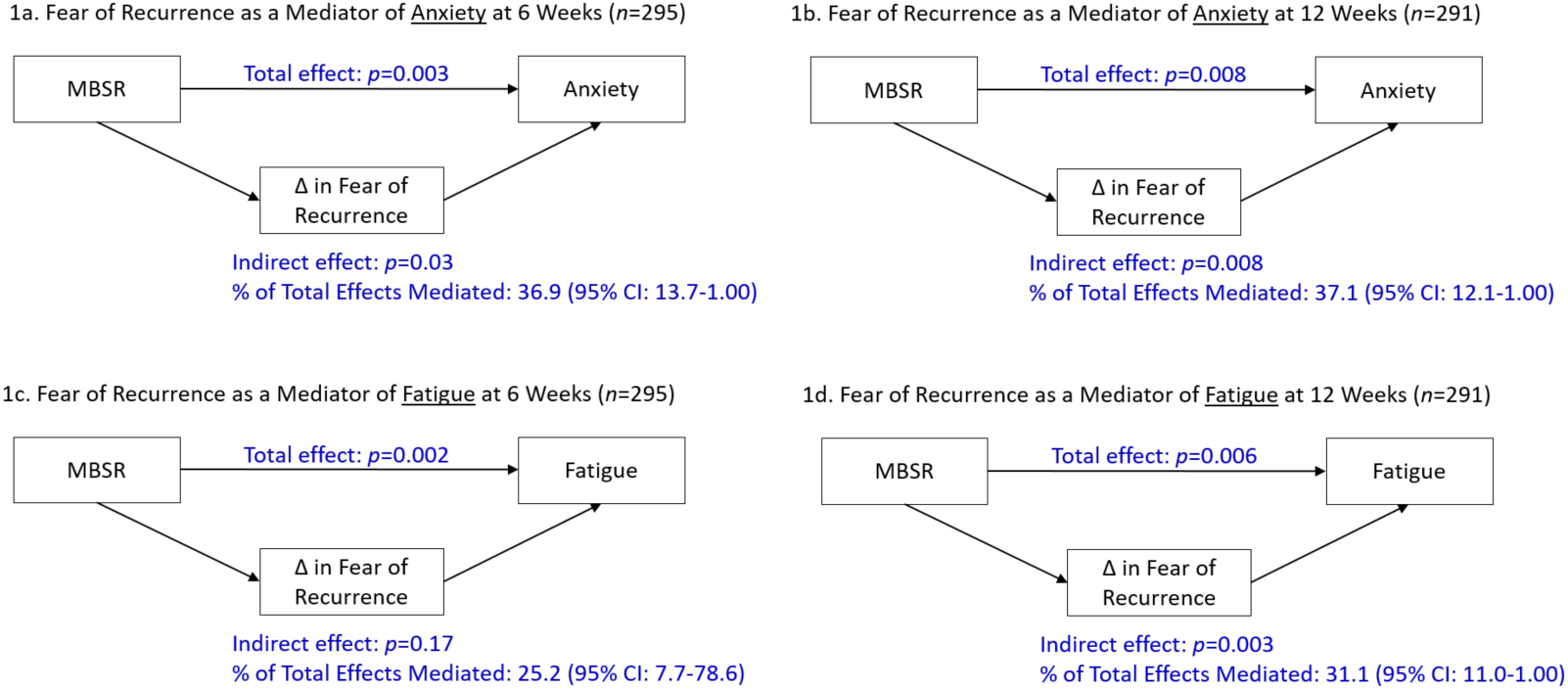
Plots of total and indirect effects of fear of recurrence as a potential mediator of state anxiety at 6 weeks (1a) and 12 weeks (1b), and fear of recurrence as a potential mediator of fatigue at 6 weeks (1c) and 12 weeks (1d).

### Perceived Stress as Mediator

Results were relatively similar when changes in perceived stress associated with the MBSR(BC) program (rather than changes in fear of recurrence) were examined in relation to levels of anxiety and fatigue at 6 and 12-weeks in a mediation analysis. Specifically, reduction in perceived stress mediated the relationship between MBSR(BC) and state anxiety at 6 and 12-weeks (*p*=0.003 and *p*=0.02, respectively) (Figure 2, top half). Of note, the reduction in perceived stress associated with the MBSR(BC) program explained 67% of the total effects of MBSR(BC) on lower levels of state anxiety at 6-weeks. Changes (reductions) in perceived stress associated with MBSR(BC) also mediated reductions in fatigue at 6 and 12-weeks (*p*=0.009 and *p*=0.04, respectively) (Figure 2, bottom half). Thus, there was substantial overlap between MBSR(BC)-induced changes in fear of recurrence and changes in perceived stress mediating the relationships between MBSR(BC) and both state anxiety and fatigue at 6- and 12-weeks. Changes in mindfulness (CAMS-R) as a potential mediator were not associated with MBSR and its effects on anxiety or fatigue at either 6 or 12 weeks (*p* > 0.90 for all 4 models).

**Figure 2.**
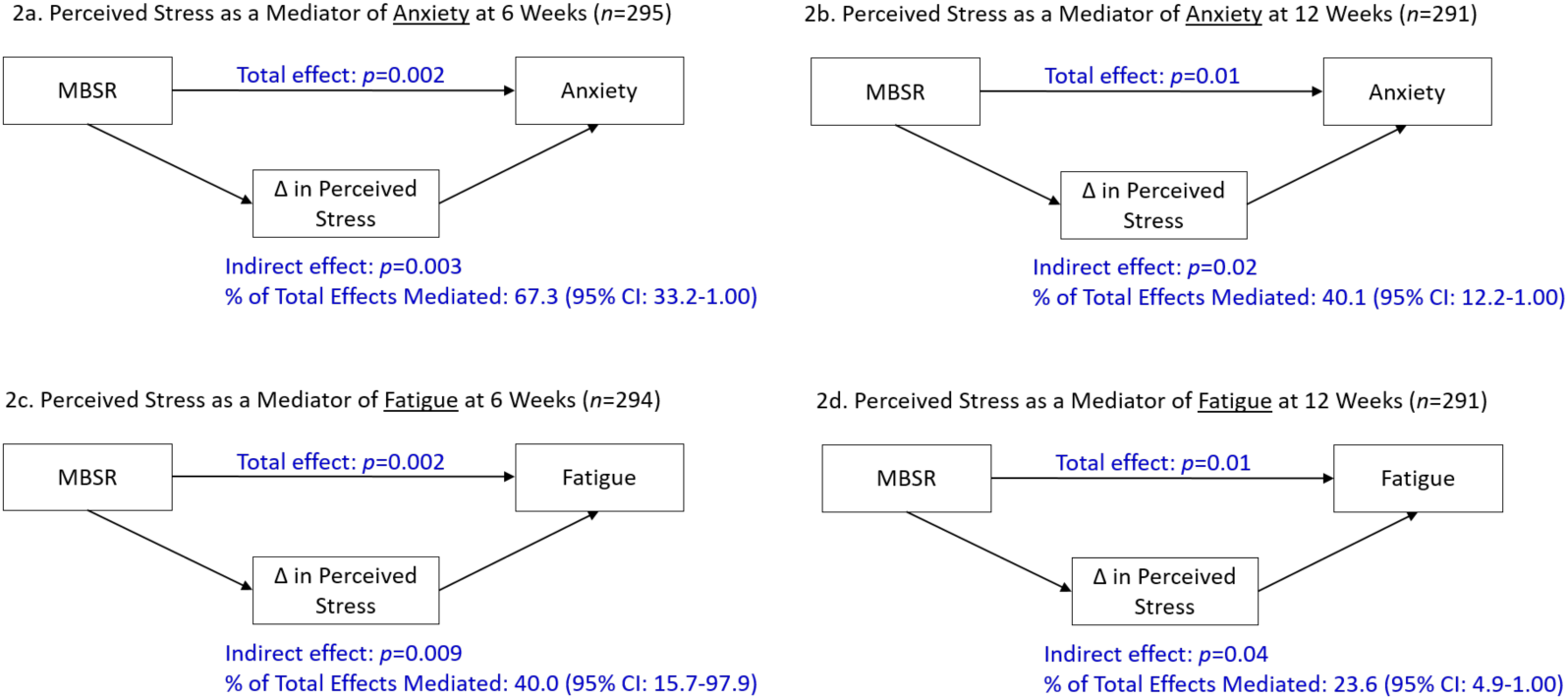
Plots of total and indirect effects of perceived stress as a potential mediator of state anxiety at 6 weeks (2a) and 12 weeks (2b), and perceived stress as a potential mediator of fatigue at 6 weeks (2c) and 12 weeks (2d).

### Subgroup Analyses

At baseline, state anxiety was strongly correlated with fatigue (*r*=0.53, *p*<0.0001), and MBSR(BC) participants presented with higher baseline state anxiety values than UC participants (Table 1). Thus, subgroup analyses were conducted to examine the total effects of MBSR(BC) and mediating effects of fear of recurrence and perceived stress on 6- and 12-week outcome scores (anxiety and fatigue) among trial participants with baseline levels of anxiety and fatigue that were above the median (*n*=111).

As seen in Table 3, a “large” total effect was observed for MBSR(BC) in relation to lower levels of fatigue at 6-weeks (effect size = 0.70, *p*=0.0002) among participants who presented with high levels of anxiety and fatigue at study entry. “Medium” effect sizes were generally observed with respect to MBSR(BC) and lower levels of anxiety, fear of recurrence, and perceived stress at both 6 and 12-weeks. Only *fear of recurrence* was selected as a mediator in the subgroup analysis due to strong effects in the overall sample. In mediational analyses, there was an indication of reductions in fear of recurrence mediating the relationship between MBSR(BC) and state anxiety at 6 and 12-weeks (*p*=0.09 and *p*=0.01) among trial participants with high anxiety and fatigue at baseline (Figure 3, top half). However, there was little to no evidence of change in fear of recurrence mediating the relationship between MBSR(BC) and fatigue at 6 and 12-weeks among trial participants with high anxiety and fatigue at baseline (Figure 3, bottom half). Thus, we did not find evidence that fear of recurrence was a stronger mediator of the relationships between MBSR(BC) and state anxiety and fatigue at 6 and 12-weeks among the subset of participants who presented with high levels of anxiety and fatigue at study entry.

**Table 3.**
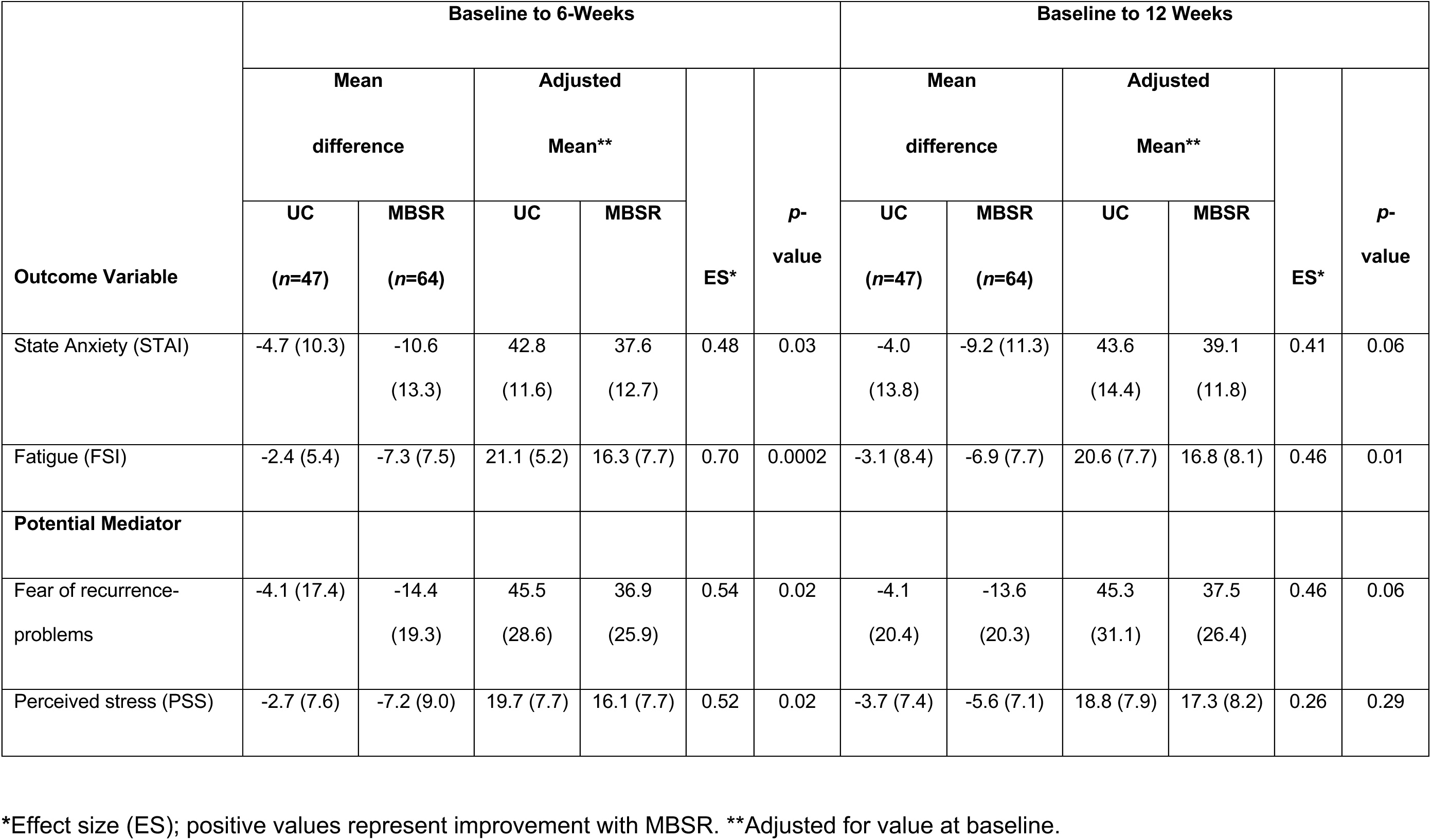
Main Effects of MBSR on Outcome and Mediating Variables at 6- and 12-Weeks among Participants who Presented with High Levels of Anxiety and Fatigue at Study Entry.

| Outcome Variable | Baseline to 6-Weeks |  |  |  |  |  | Baseline to 12 Weeks |  |  |  |  |  |
| --- | --- | --- | --- | --- | --- | --- | --- | --- | --- | --- | --- | --- |
|  | Mean difference |  | Adjusted Mean** |  | ES* | p-value | Mean difference |  | Adjusted Mean** |  | ES* | p-value |
|  | UC<br>(n=47) | MBSR<br>(n=64) | UC | MBSR |  |  | UC<br>(n=47) | MBSR<br>(n=64) | UC | MBSR |  |  |
| State Anxiety (STAI) | -4.7 (10.3) | -10.6<br>(13.3) | 42.8<br>(11.6) | 37.6<br>(12.7) | 0.48 | 0.03 | -4.0<br>(13.8) | -9.2 (11.3) | 43.6<br>(14.4) | 39.1<br>(11.8) | 0.41 | 0.06 |
| Fatigue (FSI) | -2.4 (5.4) | -7.3 (7.5) | 21.1 (5.2) | 16.3 (7.7) | 0.70 | 0.0002 | -3.1 (8.4) | -6.9 (7.7) | 20.6 (7.7) | 16.8 (8.1) | 0.46 | 0.01 |
| Potential Mediator |  |  |  |  |  |  |  |  |  |  |  |  |
| Fear of recurrence-problems | -4.1 (17.4) | -14.4<br>(19.3) | 45.5<br>(28.6) | 36.9<br>(25.9) | 0.54 | 0.02 | -4.1<br>(20.4) | -13.6<br>(20.3) | 45.3<br>(31.1) | 37.5<br>(26.4) | 0.46 | 0.06 |
| Perceived stress (PSS) | -2.7 (7.6) | -7.2 (9.0) | 19.7 (7.7) | 16.1 (7.7) | 0.52 | 0.02 | -3.7 (7.4) | -5.6 (7.1) | 18.8 (7.9) | 17.3 (8.2) | 0.26 | 0.29 |

\*Effect size (ES); positive values represent improvement with MBSR. \*\*Adjusted for value at baseline.

**Figure 3.**
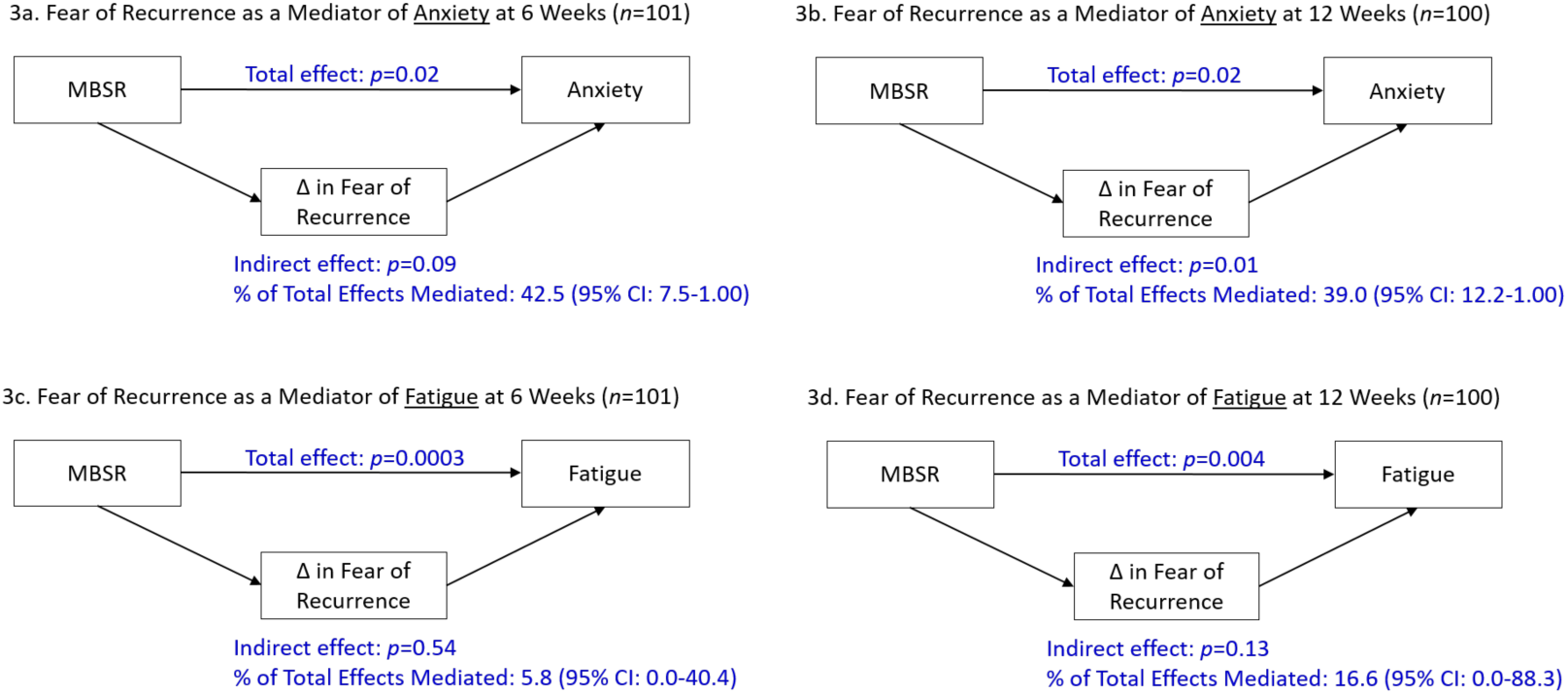
Plots of total and indirect effects of fear of recurrence as a potential mediator of state anxiety at 6 weeks (3a) and 12 weeks (3b), and fear of recurrence as a potential mediator of fatigue at 6 weeks (3c) and 12 weeks (3d). The analysis is restricted to participants who presented with levels of anxiety and fatigue above the median at study entry.

In summary the results of this current study examining BCS (n=322) in transition post-treatment suggest that the observed reductions in state anxiety and fatigue in the MBSR(BC) group are mediated by reductions in fear of recurrence and perceived stress.

## DISCUSSION

As part of a larger R01 clinical trial, results from these mediation analyses provide new knowledge and evidence identifying potential mechanisms of action for the MBSR(BC) intervention among breast cancer survivors (BCS) who are in transition off treatment. After initial analyses of early intervention effects of MBSR(BC) (baseline to 6 weeks), results pointed to fear of recurrence overall and fear of recurrence problems, and to a lesser extent perceived stress, as potential mediators of MBSR(BC) effects. Mindfulness was not identified to be a potential mediator from baseline to 6 weeks. Late intervention effects from baseline to 12 weeks were observed in fear of recurrence and trends toward perceived stress and mindfulness. These findings indicate that some mechanisms of action for MBSR(BC) may occur through the internal, cognitive perspectives of one’s affect (fear of recurrence/future uncertainties, self-perception of stress) to successfully reduce anxiety and fatigue. This evidence contributes to the scientific knowledge of MBSR by identifying specific cognitive mechanisms of action through which the MBSR(BC) program may positively influence health outcomes among BCS.

### Mindfulness as a mediator

In a systematic review of 19 mind-body interventions, 4 studies incorporated elements of mindfulness, however, mindfulness did not significantly reduce fear of cancer recurrence (Hall et al., 2018). Hall and colleagues (2018) addressed the current discussion on what mechanisms of MBSR may improve health outcomes, with mindfulness possibly not playing a central role in the mechanism pathway of fear and stress to improve anxiety and fatigue. In this study, mindfulness effects increased between 6 and 12 weeks, but never reached statistical significance, suggesting that cognitive changes specific to mindfulness may be a gradual process. Longer mindfulness meditation practice has been associated with improved optimism (C. A. Lengacher et al., 2009), improved psychological well-being (Baer et al., 2008; Josefsson, Larsman, Broberg, & Lundh, 2011), and reduced post-traumatic stress symptoms of avoidance (Bränström, Kvillemo, & Moskowitz, 2012). These findings suggest that the effectiveness of the MBSR(BC) intervention may require a long-term commitment to practice attaining self-regulation of reactions to stress for psychological health improvements. Furthermore, in a waitlist control study of women with cancer, mindfulness did not mediate the effect of the MBSR intervention on depression (L.E. Labelle, Campbell, & Carlson, 2010). Mindfulness therefore may not be the core mechanism determining MSBR’s effects on depression and fatigue (a correlated symptom of depression).

### Stress and fear as mediators

Perceived stress and fear of recurrence were found to mediate the positive effects of MBSR(BC) on psychological health outcomes of anxiety and fatigue. Perceived stress mediated the effects of MBSR(BC) on state anxiety and fatigue. Prior to this study, reduced stress and fear have been evaluated as outcome measures of mindfulness (non-MBSR) interventions in non-cancer populations (Kim, Lee, Kim, Choi, & Lee, 2016; O’Bryan, Luberto, Kraemer, & McLeish, 2018), as well as MBSR interventions in cancer populations (Butow et al., 2017; Cheli et al., 2019; Compen et al., 2018; Crane-Okada et al., 2012; C.A. Lengacher et al., 2011; C. A. Lengacher et al., 2009; C. A. Lengacher et al., 2016). Examining stress as an underlying physiological-mechanistic process of how the MBSR intervention works, have largely remained unexplored. This study provides further evidence that “fear” and “perceived stress” are cognitive mechanisms for symptom improvement in anxiety and fatigue. These results further validate “fear of recurrence” as a mediator of the MBSR(BC) intervention on symptom improvement (Lengacher et al, 2014).

### Mediation effects on fatigue

While perceived stress was found to be a mediator of fatigue, “fear of recurrence” did not mediate fatigue. Furthermore, despite a large effect size in the subgroup analysis for MBSR(BC) reducing fatigue (0.7), the effects explained by the mediators were minimal. MBSR(BC) seems to be most effective among BCS experiencing high anxiety and fatigue (C. A. Lengacher, Reich, Paterson, Ramesar, Park, Alinat, Johnson-Mallard, Moscoso, Budhrani-Shani, et al., 2016), i.e. individuals who may benefit most from the intervention. Since fatigue is one of the most distressing symptoms identified for BCS, understanding how the MBSR(BC) reduces fatigue is vital to future research. One could hypothesize that if MBSR(BC) self regulates reactions to emotional distress, this in turn may decrease emotional fatigue. Future research may also test if decreased rumination may be the process through which fatigue is decreased.

### Alternative cognitive mechanisms not tested

Self-compassion as part of the MBSR program is identified to be a vital component that may more narrowly focus on reducing suffering (Germer, 2009; Goldstein & Kornfield, 2001). Self-compassion requires one to respond to distressing and painful thoughts with self-kindness, thus reducing the perceived severity of the situation, and allowing one to maintain mindful awareness and attention to unpleasant experiences (Baer, Lykins, & Peters, 2012). Enhanced self-compassion may provide improvements in psychological well-being through self-regulatory emotional and cognitive control, thereby reducing anxiety and stress. Another cognitive mechanism that may improve health outcomes is “cognitive flexibility”: the viewing of thoughts, feelings and experiences without avoiding or ruminating on their negative qualities (Hayes, Luoma, Bond, Masuda, & Lillis, 2006). Psychological flexibility was found to be a significant mediator on stress and mood among cancer patients (Hayes et al., 2006; Laura Elizabeth Labelle, 2012). As part of the MBSR(BC) program, to achieve maximum benefits, practice and proficiency in self-regulating reactions to distress through increased attention and awareness remain critical elements that may contribute to increased psychological or cognitive flexibility.

Future research may also examine cognitive and neurobiological mechanisms underlying self-compassion in relation to MBSR(BC) as an intervention to improve well-being. In the same vein, MBSR has shown promising results in reducing rumination in cancer populations (Campbell, Labelle, Bacon, Faris, & Carlson, 2012; Jain et al., 2007), but this has yet to be examined as a possible mechanism of MBSR in lowering stress and anxiety. A limitation of this current study is that self-reported fear and stress are naturally intertwined with self-reported anxiety, and therefore require further investigations utilizing objective measures of how these cognitive mediators are functionally changing during and after the MBSR(BC) intervention period.

## CONCLUSION

This study contributes to the advancement of new knowledge and the scientific understanding of how the MBSR(BC) program may work through the cognitive mechanisms used by participants as part of the intervention. Specifically, the effectiveness of MBSR(BC) is believed to occur by participants incorporating greater emotional and cognitive control through self-regulation of reactions to internal and external experiences. With a paucity of researchers investigating these mechanisms, this current study provides support for cognitive mechanisms being central to the success of the MBSR(BC) intervention. Ultimately, the goal of this work is to increase the QOL and improve distressing symptoms among BCS transitioning off treatment. The major benefits of performing the mediation analyses for this study are to advance clinical treatment research for cancer survivors thus providing evidence for intervention components (Moyer et al., 2012). Testing the mechanisms of action advances intervention research by “optimizing” the therapeutic active benefits and tailoring interventions more specifically for BCS (Alan E. Kazdin, 2007). Results of this mediation analyses may contribute to advancement of symptom science and theory development (Alan E. Kazdin, 2007), through a more meaningful understanding of mindfulness (K. W. Brown, R. M. Ryan, & J. D. Creswell, 2007).

We postulated that to achieve the maximum benefit from the MBSR(BC) program, “practice and proficiency” in mindfulness are critically important elements through reducing fear of recurrence and perceived stress. MBSR(BC) may result in cognitive adaptations that in turn result in reduced physical and psychological symptoms and increased well-being. To advance the science of empirical investigations of intervention strategies, it is essential within larger clinical trials to validate MBSR intervention mechanisms that are specifically tailored for cancer survivors.

## Data Availability

Data will be archived in accordance with the NIH Data Sharing Plan, a national data repository.

## REFERENCES

Baer, R. A., Lykins, E., & Peters, J. R. (2012). Mindfulness and self-compassion as predictors of psychological wellbeing in long-term meditators and matched nonmeditators. The Journal of Positive Psychology, 7(3), 230–238. doi:10.1080/17439760.2012.674548

Baer, R. A., Smith, G. T., Hopkins, J., Krietemeyer, J., & Toney, L. (2006). Using self-report assessment methods to explore facets of mindfulness. Assessment, 13(1), 27–45. doi:10.1177/1073191105283504

Baer, R. A., Smith, G. T., Lykins, E., Button, D., Krietemeyer, J., Sauer, S.,... Williams, J. M. (2008). Construct validity of the five facet mindfulness questionnaire in meditating and nonmeditating samples. Assessment, 15(3), 329–342. doi:10.1177/1073191107313003

Bishop, S. R. (2002). What do we really know about mindfulness-based stress reduction? Psychosomatic Medicine 64(1), 71–83. Retrieved from http://www.ncbi.nlm.nih.gov/entrez/query.fcgi?cmd=Retrieve&db=PubMed&dopt=Citation&list_uids=11818588

Bishop, S. R., Lau, M., Shapiro, S., Carlson, L., Anderson, N. D., Carmody, J.,... Devins, G. (2004). Mindfulness: A proposed operational definition. Clinical Psychology Science and Practice, 11(3), 230–241. doi:10.1093/clipsy/bph077

Boyle, C. C., Stanton, A. L., Ganz, P. A., Crespi, C. M., & Bower, J. E. (2017). Improvements in emotion regulation following mindfulness meditation: Effects on depressive symptoms and perceived stress in younger breast cancer survivors. Journal of Consulting and Clinical Psychology, 85(4), 397.

Branstrom, R., Kvillemo, P., Brandberg, Y., & Moskowitz, J. T. (2010). Self-report mindfulness as a mediator of psychological well-being in a stress reduction intervention for cancer patients-A randomized study. Annals of Behavioral Medicine 39(2), 151–161. doi:10.1007/s12160-010-9168-6

Bränström, R., Kvillemo, P., & Moskowitz, J. T. (2012). A randomized study of the effects of mindfulness training on psychological well-being and symptoms of stress in patients treated for cancer at 6-month follow-up. International journal of behavioral medicine, 19(4), 535–542.

Brown, K. W., & Cordon, S. (2009). Toward a Phenomenology of Mindfulness: Subjective Experience and Emotional Correlates. In F. Didonna (Ed.), Clinical Handbook of Mindfulness. New York: Springer Science.

Brown, K. W., Ryan, R. A., & Creswell, J. D. (2007). Mindfulness: Theoretical foundations and evidence for its salutary effects. Psychological Inquiry, 18(4), 211–237. Retrieved from <Go to ISI>://000250865000001

Brown, K. W., Ryan, R. M., & Creswell, J. D. (2007). Mindfulness: Theoretical Foundations and Evidence for its Salutary Effects. Psychological Inquiry, 18(4), 211–237. doi:10.1080/10478400701598298

Butow, P. N., Turner, J., Gilchrist, J., Sharpe, L., Smith, A. B., Fardell, J. E.,... Gebski, V. J. (2017). Randomized trial of ConquerFear: a novel, theoretically based psychosocial intervention for fear of cancer recurrence. Journal of Clinical Oncology, 35(36), 4066–4077.

Campbell, T. S., Labelle, L. E., Bacon, S. L., Faris, P., & Carlson, L. E. (2012). Impact of mindfulness-based stress reduction (MBSR) on attention, rumination and resting blood pressure in women with cancer: a waitlist-controlled study. Journal of Behavioral Medicine, 35(3), 262–271.

Carlson, L. E., Doll, R., Stephen, J., Faris, P., Tamagawa, R., Drysdale, E., & Speca, M. (2013). Randomized controlled trial of mindfulness-based cancer recovery versus supportive expressive group therapy for distressed survivors of breast cancer (MINDSET). Journal of Clinical Oncology, 31(25), 3119–3126. doi:10.1200/jco.2012.47.5210

Carpenter, J. S., & Andrykowski, M. A. (1998). Psychometric evaluation of the Pittsburgh Sleep Quality Index. Journal of Psychosomatic Research, 45(1 Spec No), 5–13. Retrieved from http://www.ncbi.nlm.nih.gov/entrez/query.fcgi?cmd=Retrieve&db=PubMed&dopt=Citation&list_uids=9720850

Cheli, S., Caligiani, L., Martella, F., De Bartolo, P., Mancini, F., & Fioretto, L. (2019). Mindfulness and metacognition in facing with fear of recurrence: A proof-of-concept study with breast-cancer women. Psycho-oncology, 28(3), 600–606.

Cohen, S., Kamarck, T., & Mermelstein, R. (1983). A global measure of perceived stress. Journal of Health and Social Behavior, 24(4), 385–396. Retrieved from http://www.ncbi.nlm.nih.gov/pubmed/6668417

Compen, F., Bisseling, E., Schellekens, M., Donders, R., Carlson, L., van der Lee, M., & Speckens, A. (2018). Face-to-face and internet-based mindfulness-based cognitive therapy compared with treatment as usual in reducing psychological distress in patients with cancer: a multicenter randomized controlled trial. Journal of Clinical Oncology, 36(23), 2413–2421.

Crane-Okada, R., Kiger, H., Sugerman, F., Uman, G. C., Shapiro, S. L., Wyman-McGinty, W., & Anderson, N. L. (2012). Mindful movement program for older breast cancer survivors: a pilot study. Cancer Nursing, 35(4), E1–E13.

DeSantis, C. E., Ma, J., Gaudet, M. M., Newman, L. A., Miller, K. D., Goding Sauer, A.,... Siegel, R. L. (2019). Breast cancer statistics, 2019. CA: A Cancer Journal for Clinicians, 69(6), 438–451.

Eisenberg, S. A., Kurita, K., Taylor-Ford, M., Agus, D. B., Gross, M. E., & Meyerowitz, B. E. (2015). Intolerance of uncertainty, cognitive complaints, and cancer-related distress in prostate cancer survivors. Psycho-Oncology, 24(2), 228–235.

Feldman, G. (2007). Cognitive and behavioral therapies for depression: Overview, new directions, and practical recommendations for dissemination. Psychiatric Clinics of North America, 30(1), 39–50. doi:S0193-953X(06)00103-1 {pii} 10.1016/j.psc.2006.12.001

Feldman, G., Hayes, A., Kumar, S., Greeson, J., & Laurenceau, J. (2007). Mindfulness and emotion regulation: The development and initial validation of the Cognitive and Affective Mindfulness Scale-Revised (CAMS-R). Journal of Psychopathology and Behavioral Assessment, 29(3), 177–190. doi:DOI 10.1007/s10862-006-9035-8

Garland, S. N., Carlson, L. E., Stephens, A. J., Antle, M. C., Samuels, C., & Campbell, T. S. (2014). Mindfulness-based stress reduction compared with cognitive behavioral therapy for the treatment of insomnia comorbid with cancer: a randomized, partially blinded, noninferiority trial. Journal of Clinical Oncology, 32(5), 449–457. doi:10.1200/jco.2012.47.7265

Germer, C. K. (2009). Mindful Path to Self-Compassion Freeing Yourself from Destructive Thoughts and Emotions. New York: Guilford Publications, Inc.

Goldstein, J., & Kornfield, J. (2001). Seeking the heart of wisdom: The path of insight meditation. Boston, MA: Shambhala Classics

Hall, D. L., Luberto, C. M., Philpotts, L. L., Song, R., Park, E. R., & Yeh, G. Y. (2018). Mind-body interventions for fear of cancer recurrence: A systematic review and meta-analysis. Psycho-oncology, 27(11), 2546–2558.

Hamilton, N. A., Kitzman, H., & Guyotte, S. (2006). Enhancing health and emotion: Mindfulness as a missing link between cognitive therapy and positive psychology. Journal of Cognitive Psychotherapy, 20(2), 123–134.

Hann, D. M., Jacobsen, P., Martin, S., Azzarello, L., & Greenberg, H. (1998). Fatigue and quality of life following radiotherapy for breast cancer: a comparative study. J Clin Psychol Med Settings, 5, 19–33.

Hayes, S., Luoma, J., Bond, F., Masuda, A., & Lillis, J. (2006). Behaviour research and therapy. Behaviour Research and Therapy, 44, 1–25.

Hicks, R., & Tingley, D. (2011). Causal mediation analysis. Stata Journal, 11(4), 605.

Imai, K., Keele, L., & Tingley, D. (2010). A general approach to causal mediation analysis. Psychological Methods, 15(4), 309–334. doi:10.1037/a0020761

Jain, S., Shapiro, S. L., Swanick, S., Roesch, S. C., Mills, P. J., Bell, I., & Schwartz, G. E. (2007). A randomized controlled trial of mindfulness meditation versus relaxation training: Effects on distress, positive states of mind, rumination, and distraction. Annuals of Behavioral Medicine, 33(1), 11–21. doi:10.1207/s15324796abm3301_2

Johns, S. A., Brown, L. F., Beck-Coon, K., Monahan, P. O., Tong, Y., & Kroenke, K. (2015). Randomized controlled pilot study of mindfulness-based stress reduction for persistently fatigued cancer survivors. Psychooncology, 24(8), 885–893. doi:10.1002/pon.3648

Josefsson, T., Larsman, P., Broberg, A., & Lundh, L.-G. (2011). Self-Reported Mindfulness Mediates the Relation Between Meditation Experience and Psychological Well-Being. Mindfulness, 2(1), 49–58. doi:10.1007/s12671-011-0042-9

Kabat-Zinn, J. (1990). Full-catastrophe living: Using the wisdom of your body and mind to face stress, pain and illness. New York, NY: Bantam Doubleday Dell Publishing.

Kabat-Zinn, J. (2003). Mindfulness-based interventions in context: Past, present, and future. Clinical Psychology-Science and Practice, 10(2), 144–156. doi:DOI 10.1093/clipsy/bpg016

Kabat-Zinn, J., Lipworth, L., & Burney, R. (1985). The clinical use of mindfulness meditation for the self-regulation of chronic pain. Journal of Behavioral Medicine, 8(2), 163–190. Retrieved from http://www.ncbi.nlm.nih.gov/entrez/query.fcgi?cmd=Retrieve&db=PubMed&dopt=Citation&list_uids=3897551

Kabat-Zinn, J., Massion, A. O., Kristeller, J., Peterson, L. G., Fletcher, K. E., Pbert, L.,... Santorelli, S. F. (1992). Effectiveness of a meditation-based stress reduction program in the treatment of anxiety disorders. American Journal of Psychiatry, 149(7), 936–943. Retrieved from http://www.ncbi.nlm.nih.gov/pubmed/1609875

Kazdin, A. E. (2006). Mechanisms of Change in Psychotherapy: Advances, Breakthroughs, and Cutting-Edge Research (Do Not Yet Exist).

Kazdin, A. E. (2007). Mediators and Mechanisms of Change in Psychotherapy Research. Annual Review of Clinical Psychology, 3(1), 1. Retrieved from http://ezproxy.lib.usf.edu/login?url=http://search.ebscohost.com/login.aspx?direct=true&db=edb&AN=53344843&site=eds-live

Keller, S., Bann, C. M., Dodd, S. L., Schein, J., Mendoza, T. R., & Cleeland, C. S. (2004). Validity of the brief pain inventory for use in documenting the outcomes of patients with noncancer pain. Clinical Journal of Pain, 20(5), 309–318. Retrieved from http://www.ncbi.nlm.nih.gov/entrez/query.fcgi?cmd=Retrieve&db=PubMed&dopt=Citation&list_uids=15322437

Kim, M. K., Lee, K. S., Kim, B., Choi, T. K., & Lee, S.-H. (2016). Impact of mindfulness-based cognitive therapy on intolerance of uncertainty in patients with panic disorder. Psychiatry investigation, 13(2), 196.

Labelle, L. E. (2012). How does mindfulness-based stress reduction (mbsr) improve psychological functioning in cancer patients?

Labelle, L. E., Campbell, T. S., & Carlson, L. E. (2010). Mindfulness-based stress reduction in oncology: Evaluating mindfulness and rumination as mediators of change in depressive symptoms. Mindfulness, 1, 28–40. doi:10.1007/s12671-010-0005-6

Labelle, L. E., Lawlor-Savage, L., Campbell, T. S., Faris, P., & Carlson, L. E. (2015). Does self-report mindfulness mediate the effect of Mindfulness-Based Stress Reduction (MBSR) on spirituality and posttraumatic growth in cancer patients? The Journal of Positive Psychology, 10(2), 153–166.

Laurenceau, J.-P., Hayes, A. M., & Feldman, G. C. (2007). Some methodological and statistical issues in the study of change processes in psychotherapy. Clinical Psychology Review, 27(6), 682–695.

Lebel, S., Maheu, C., Lefebvre, M., Secord, S., Courbasson, C., Singh, M.,... Fung, M. F. K. (2014). Addressing fear of cancer recurrence among women with cancer: a feasibility and preliminary outcome study. Journal of Cancer Survivorship, 8(3), 485–496.

Lee-Jones, C., Humphris, G., Dixon, R., & Hatcher, M. B. (1997). Fear of cancer recurrence--a literature review and proposed cognitive formulation to explain exacerbation of recurrence fears. Psycho-Oncology, 6(2), 95–105. Retrieved from http://www.ncbi.nlm.nih.gov/pubmed/9205967

Lengacher, C. A., Johnson-Mallard, V., Barta, M., Fitzgerald, S., Moscoso, M. S., Post-White, J.,... Kip, K. E. (2011). Feasibility of a mindfulness-based stress reduction program for early-stage breast cancer survivors. Journal of Holistic Nursing, 29(2), 107–117. doi:10.1177/0898010110385938

Lengacher, C. A., Johnson-Mallard, V., Post-White, J., Moscoso, M. S., Jacobsen, P. B., Klein, T. W.,... Kip, K. E. (2009). Randomized controlled trial of mindfulness-based stress reduction (MBSR) for survivors of breast cancer. Psycho-Oncology, 18(12), 1261–1272. doi:10.1002/pon.1529

Lengacher, C. A., Reich, R., Post-White, J., Moscoso, M., Shelton, M., Barta, M.,... Budhrani, P. (2012). Mindfulness based stress reduction in post-treatment breast cancer patients: An examination of symptoms and symptom clusters. Journal of Behavioral Medicine, 35(1), 86–94. doi:10.1007/s10865-011-9346-4

Lengacher, C. A., Reich, R. R., Paterson, C. L., Jim, H. S., Ramesar, S., Alinat, C. B.,... Kip, K. E. (2015). The effects of mindfulness-based stress reduction on objective and subjective sleep parameters in women with breast cancer: A randomized controlled trial. Psycho-Oncology, 24(4), 424–432. doi:10.1002/pon.3603

Lengacher, C. A., Reich, R. R., Paterson, C. L., Ramesar, S., Park, J. Y., Alinat, C.,... Kip, K. E. (2016). Examination of Broad Symptom Improvement Resulting From Mindfulness-Based Stress Reduction in Breast Cancer Survivors: A Randomized Controlled Trial. Journal of Clinical Oncology, 34(24), 2827–2834. doi:10.1200/jco.2015.65.7874

Lengacher, C. A., Reich, R. R., Paterson, C. L., Ramesar, S., Park, J. Y., Alinat, C. B.,... Kip, K. E. (2016). Examination of broad symptom improvement due to Mindfulness-Based Stress Reduction for breast cancer survivors: A randomized controlled trial. Journal of Clinical Oncology, 37(24), 2827–2834. doi:10.1200/JCO.2015.65.7874

Lengacher, C. A., Shelton, M. M., Reich, R. R., Barta, M. K., Johnson-Mallard, V., Moscoso, M. S.,... Kip, K. E. (2014). Mindfulness based stress reduction (MBSR(BC)) in breast cancer: Evaluating fear of recurrence (FOR) as a mediator of psychological and physical symptoms in a randomized control trial (RCT). Journal of Behavioral Medicine 37(2), 185–195. doi:10.1007/s10865-012-9473-6

Moyer, A., Goldenberg, M., Hall, M. A., Knapp-Oliver, S. K., Sohl, S. J., Sarma, E. A., & Schneider, S. (2012). Mediators of change in psychosocial interventions for cancer patients: A systematic review. Behavioral Medicine, 38(3), 90–114.

Mutsaers, B., Jones, G., Rutkowski, N., Tomei, C., Leclair, C. S., Petricone-Westwood, D.,... Lebel, S. (2016). When fear of cancer recurrence becomes a clinical issue: a qualitative analysis of features associated with clinical fear of cancer recurrence. Supportive Care in Cancer, 24(10), 4207–4218.

O’Bryan, E. M., Luberto, C. M., Kraemer, K. M., & McLeish, A. C. (2018). An examination of mindfulness skills in terms of affect tolerance among individuals with elevated levels of health anxiety. Anxiety, Stress, & Coping, 31(6), 702–713.

Preacher, K. J., & Hayes, A. F. (2004). SPSS and SAS procedures for estimating indirect effects in simple mediation models. Behav Res Methods Instrum Comput, 36(4), 717–731.

Radloff, L. (1977). The CES-D scale: A self-report depression scale for researching the general population. Application of Psychological Measures, 1(3), 385–401.

Reich, R. R., Lengacher, C. A., Alinat, C. B., Kip, K. E., Paterson, C., Ramesar, S.,... Moscoso, M. (2017). Mindfulness-based stress reduction in post-treatment breast cancer patients: Immediate and sustained effects across multiple symptom clusters. Journal of Pain and Symptom Management, 53(1), 85–95. doi:10.1016/j.jpainsymman.2016.08.005

Society, A. C. (2020). Survival Rates for Breast Cancer Retrieved from https://www.cancer.org/cancer/breast-cancer/understanding-a-breast-cancer-diagnosis/breast-cancer-survival-rates.htmlBaer, R. A. (2003). Mindfulness training as a clinical intervention: A conceptual and empirical review. *Clinical Psychology: Science and Practice*, 10(2), 125–143. doi:10.1093/clipsy/bpg015

Society, A. C. (2019). Cancer Facts & Figures. In A. C. Society (Ed.). Atlanta.

Spielberger, C., Gorsuch, R., & Luschene, R. (1983). Manual for the State-Trait Anxiety Inventory. Palo Alto, CA: Consulting Psychologists.

Vickberg, S. M. (2003). The Concerns About Recurrence Scale (CARS): a systematic measure of women’s fears about the possibility of breast cancer recurrence. Annuals of Behavioral Medicine, 25(1), 16–24. Retrieved from http://www.ncbi.nlm.nih.gov/pubmed/12581932

Victorson, D., Du, H. Y., Hankin, V., King, K., McCurdy, M., Pruitt, J.,... Brendler, C. (2012). Mindfulness based stress reduction decreases fear of progression over time for men with prostate cancer on active surveillance: Results from a randomized clinical trail. Paper presented at the Journal of Urology. <Go to ISI>://WOS:000302912500384

Ware, J. H., Kosinski, M., & Keller, M. (1994). SF-36 physical and mental health summary scales, a user’s manual (Second ed.). Boston: The Health Institute.

Xu, W., Zhou, Y., Fu, Z., & Rodriguez, M. (2017). Relationships between dispositional mindfulness, self-acceptance, perceived stress, and psychological symptoms in advanced gastrointestinal cancer patients. Psycho-oncology, 26(12), 2157–2161.

Zainal, N. Z., Booth, S., & Huppert, F. A. (2013). The efficacy of mindfulness-based stress reduction on mental health of breast cancer patients: A meta-analysis. Psycho-Oncology, 22(7), 1457–1465. doi:10.1002/pon.3171

